# Diagnostic performance of rK28- and rK39-based rapid diagnostic tests for Visceral Leishmaniasis in Kenya: Implications for the national diagnostic algorithm

**DOI:** 10.64898/2026.11.13.26368071

**Authors:** Damaris Matoke-Muhia, Mercy Tuluso, Bryson B. Kimemia, Benard Osero, Luna Kamau, Joseph M. Ndung’u, Robert Ofwete, Kelvin Thiong’o, Dawn Maranga

## Abstract

**Background:** Visceral Leishmaniasis (VL) is a neglected tropical disease (NTD) caused by *Leishmania infantum* and *L. donovani* and transmitted by a bite of an infected female phlebotomine sandfly. The disease disproportionately affects impoverished and underserved populations, where inadequate access to healthcare contributes to delayed diagnosis, treatment, and increased morbidity and mortality. In Kenya, 11 out of the 47 counties are known to be endemic for VL. Visceral Leishmaniasis is fatal if it’s not timely diagnosed and treated. Parasitological methods remain the gold standard for diagnosing and confirming VL, but they are invasive and require specialised training and equipment. Serological tests in the form of Rapid Diagnostic Tests (RDTs) are widely used for initial screening and detection but can still report positive results for long periods after curative treatment due to the persistence of anti-VL antibodies. It is recommended that VL diagnosis follows an algorithmic approach. This study evaluated the diagnostic performance of rK28 and rK39 RDTs (Kala-azar Detect and IT Leish) in various algorithmic combinations against a composite reference standard in Kenya.

**Methodology:** We evaluated rK28 and Kala-azar Detect along with rK39 IT-Leish as index tests, and Direct Agglutination Test (DAT) as the reference standard test. Participants were recruited from seven health facilities located in V<u>L</u> endemic regions of Marsabit and Turkana counties, Kenya. Blood samples were collected, as well as patient demographic data. Treatment decisions followed the national diagnostic algorithm and health facility routine procedures. Diagnostic test performance was assessed individually, in parallel, and based on sequential algorithmic combinations. Specificity, sensitivity, balanced accuracy, and negative and positive predictive values were statistically estimated.

**Results:** A total of 398 participants were recruited for the study, with fever and splenomegaly being the most reported clinical symptoms. Individually, the rK28 kit demonstrated a sensitivity of 96%, a specificity of 86%, and a balanced accuracy (BA) of 91.2%. The rK39 IT-Leish kit showed a sensitivity of 87%, a specificity of 93%, and a BA of 90.4%, while the rK39 Kala-azar Detect showed a sensitivity of 67%, a specificity of 91%, and a BA of 79%. The rK39 IT Leish to rK28 algorithm based on negatives had the highest balanced accuracy of 90% with a sensitivity of 92% and a specificity of 88%, while the rK28 to rK39 Kala-azar detect algorithm based on positives showed the lowest balanced accuracy of 80% with a sensitivity of 65% and a specificity of 94.1%.

**Discussion:** The rK28 RDT demonstrated consistently high sensitivity (96–98%) and moderate specificity (72–86%), findings broadly consistent with reports from other East African settings. In contrast, the rK39-based tests showed a higher specificity (91–93%) but lower sensitivity (67–87%), which limits their effectiveness for elimination-focused programs that lean towards not missing a positive case. Parallel testing algorithms that combine the rK28 and rK39 RDTs improved sensitivity (up to 97.7%), whereas sequential (based on confirming negative results) approaches increased specificity at the expense of sensitivity when compared to the rK28 RDT alone. These approaches were more balanced as compared to sequential (based on confirming positive results), which increased the overall specificity but greatly impacted the sensitivity. This flexibility in combinations provides more options for programs based on risks and overall goals. The performance of the rK28 RDT gives it an advantage over the rK39 kits, especially with rK39 IT Leish kits facing stockouts. While the study faced limitations of lack of PCR data that led to the reliance on DAT as the reference standard, as well as underpowered secondary analyses, the findings point towards strong preliminary evidence that can be used to guide updating of the national algorithm, supply chain strengthening, and policy decisions that may accelerate VL elimination in the country and the region.

**Conclusion:** The findings demonstrate that diagnostic algorithms incorporating rK28 provide robust performance for VL diagnosis. These results support consideration of the rK28 diagnostic kit as a valuable component of VL diagnostic strategies, with the potential to enhance case detection and contribute to VL control and elimination efforts in Eastern Africa.

## Introduction

Visceral leishmaniasis (VL), or kala-azar, is a severe and potentially fatal disease caused by *Leishmania* parasites(1). Although around 20 species of *Leishmania* parasites cause leishmaniasis, only two (*Leishmania infantum* and *L. donovani*) are attributed to causing VL(2,3). The parasites are zoonotic and are transmitted by female phlebotomine sandflies during feeding(4). Visceral Leishmaniasis is estimated to have a global incidence rate of approximately 500,000 cases annually, resulting in approximately 50,000 deaths, with between 200 million and 400 million people at risk of contracting the disease (1,5). The eastern African region has the highest burden in the world, with the disease disproportionately affecting poverty-stricken communities with limited access to medical care and proper nutrition (6).

In Kenya, approximately 4,000 cases of VL are reported annually, with around 5 to 6 million people at risk of being infected(7,8). According to the WHO, Kenya was second only to Sudan in the number of cases reported globally in 2024(9). Out of the 47 counties in Kenya, eleven (Baringo, Garissa, Kajiado, Kitui, Isiolo, Mandera, Marsabit, Tharaka Nithi, Wajir, West Pokot, and Turkana) are known to be endemic for VL, with a case fatality rate of 0.5%(10). The Ministry of Health has also classified Samburu in the Rift Valley region as an endemic county in the latest 2021-2025 Leishmaniasis control strategy(11). These counties have experienced numerous recorded outbreaks of the disease since 2008, with the latest outbreak occurring in 2025(12). As Kenya moves towards VL elimination in line with the World Health Organisation (WHO) roadmap for NTDs 2021-2030(13), it continues to face several challenges, including political, socio-cultural, epidemiological, health system, diagnostic, and treatment access challenges. Addressing these challenges is essential(14). These challenges hinder meaningful progress towards eliminating the disease as a public health problem. Central to these efforts is access to timely and accurate diagnosis, which enables prompt treatment, strengthens surveillance, and supports effective case management in line with WHO recommendations (15).

The clinical manifestations of VL are similar to those of other febrile illnesses, such as enteric and malaria infections(16,17). This similarity, when coupled with a mortality rate of 95% to 100% in two years if left untreated,(18,19) underscores the high cost of misdiagnosis. VL is diagnosed through a combination of clinical signs and laboratory tests, which can be serological or parasitological (20). Clinical symptoms of VL include weight loss, enlargement of the spleen and liver, fever lasting more than 2 weeks, and anemia (1,21). The presence of the parasite can be determined through investigation by microscopy of samples from the lymph nodes, spleen and bone marrow aspirates, liver biopsy or the buffy coat of blood from extremities (peripheral blood) (22,23). In Kenya, the diagnostic algorithm for parasitological diagnosis is outlined in the national guidelines for the prevention, diagnosis and treatment of VL(24). While parasitological tests remain the gold standard for VL detection, the need for specialized laboratories, skilled personnel, moderate sensitivity, and the high invasiveness and discomfort associated with specimen collection remain limiting factors for this approach(25). Serological tests used in VL diagnosis include: Direct Agglutination Tests (DAT), Immunofluorescent Antibody Test (IFAT), Immunochromatographic Tests (ICT), Western blot, and Enzyme-Linked Immunosorbent Assays (ELISA) (26). In Kenya, diagnosis based on serological tests such as DAT and Immunochromatographic Tests such as the rK39 rapid diagnostic tests (RDTs) can be used to initiate treatment, only if the VL suspect case definition is strictly followed. Serological tests can still report a positive result several years after an individual was infected and was successfully treated for VL, highlighting the importance of the patient’s history during diagnosis(24). The current Ministry of Health (MoH) guidelines stipulate that new suspect cases should be initially tested with RDTs, then DAT if negative, and finally parasitological screening if the first two tests are negative. For a previous VL case, diagnosis should only be confirmed through parasitological tests(24).

Immunochromatographic RDTs, also known as lateral flow strip tests, are low-cost point-of-care tests that allow for the quick identification of disease-related analytes such as antibodies or antigens(27). The most used RDTs for the diagnosis of VL use a recombinant form of the kinesin-related, 39-amino acid residue (k-39) from *L. chagasi*, dubbed rK39, to target anti-k39 antibodies produced during a VL infection^27^. These tests can use whole blood or serum, which is a less invasive method of diagnosis as compared to the aspirates and biopsies used for parasitological diagnosis(28). In East Africa, two rk39 RDTs are used, namely: IT Leish^®^ previously manufactured by Bio-Rad (Hercules, California, United States) and then by GADx (UK), and Kalazar Detect™ by InBios (Seattle, Washington, United States)(29). Rk39 RDTs, although quick, cost-effective, stable, and simple to use, have some drawbacks. These include varied reported sensitivities and specificities ranging from 27% to 100%, as well as the inability to differentiate between present and past infections(30). The RDTs have had a lower sensitivity in East Africa (85%) as compared to the Indian subcontinent (97%), which had the highest. The East African pooled sensitivities reported are also lower than the global average of 92%. These differences have been attributed to the presence of VL antibodies in asymptomatic individuals as well as divergence of *L. donovani* strains between the regions(31). The rK39 IT Leish^®^ is also facing stockouts due to a reported halt in production(32). A new generation kit based on the K28 recombinant synthetic antigen, dubbed rK28, was developed by Blue Heron Biotechnology (Bothell, Washington, United States) to address some of the challenges of the rK39 RDTs(33,34). This synthetic antigen contains the K39 protein, the entire hydrophilic acylated surface protein B2 (HASPB2), and several repeats of the HASPB1 protein. This increases the antigen epitope density, which has already shown markedly improved performance in Sudan(33,35). These tests can use whole blood or serum, which is a less invasive method of diagnosis as compared to the aspirates and biopsies used for parasitological diagnosis(28). In East Africa, two rk39 RDTs are used, namely: DiaMED-IT Leish® by Bio-Rad (Hercules, California, United States) and Kalazar Detect™ (Seattle, Washington, United States)(29). Rk39 RDTs, although quick, cost-effective, stable, and simple to use, have some drawbacks. These include varied reported sensitivities and specificities, in some cases as low as 27% but up to 100% in others, as well as the inability to differentiate between present and past infections(30). The RDTs have had a lower pooled sensitivity in East Africa (85%) as compared to the Indian subcontinent (97%), which had the highest. The East African pooled sensitivities reported are also lower than the global average of 92%. These differences have been attributed to the presence of VL antibodies in asymptomatic individuals as well as divergence of *L. donovani* strains between the regions(31). A new generation kit based on the K28 recombinant synthetic antigen, dubbed rK28, was developed to address some of the challenges of the rK39 RDTs(33,34). This synthetic antigen contains the K39 protein, the entire hydrophilic acylated surface protein B2 (HASPB2), and several repeats of the HASPB1 protein. This increases the antigen epitope density, which has already shown a marked improvement in performance in Sudan(33,35).

This study evaluated the diagnostic performance of rK28, rK39 IT-Leish, and rK39 Kala-azar Detect, individually and in algorithmic combinations, using DAT as the reference standard. The study was conducted across seven sentinel sites in northern Kenya. The study provides essential evidence to inform national diagnostic policy, optimise RDT deployment across health system levels, and support ongoing VL elimination efforts.

## Materials and Methods

### Study Design

This was a single-arm, non-interventional, prospective diagnostic accuracy study evaluating the performance of rK28, Kala-azar Detect, and IT-Leish RDTs, individually and in algorithmic combinations. The originally pre-specified composite reference standard included DAT, PCR, and microscopy where available.

### Study Participants

Participants were consecutively enrolled as they presented to seven health facilities in Marsabit (Laisamis, Marsabit and Moyale) and Turkana (Lopiding, Kakuma, Lodwar and Namoruputh) counties in northern Kenya (**Fig 1**) between January 2023 and March 2024. The study sites included health centres, sub-county hospitals, and county referral hospitals selected based on VL burden and diagnostic capacity^33^. Eligible participants were individuals aged ≥1 year presenting with clinical signs and symptoms suggestive of VL, including fever of more than two weeks, splenomegaly, and weight loss. Individuals who tested negative for malaria and were considered VL suspects according to national guidelines were also eligible for participation. Written informed consent was obtained from all adult participants and from parents or legal guardians for minors, with assent obtained from children aged 12–17 years where applicable. Individuals with VL relapses within the previous 6–12 months, post-kala-azar dermal leishmaniasis (PKDL), or inability to provide the required 2ml of blood were excluded.

**Fig 1:**
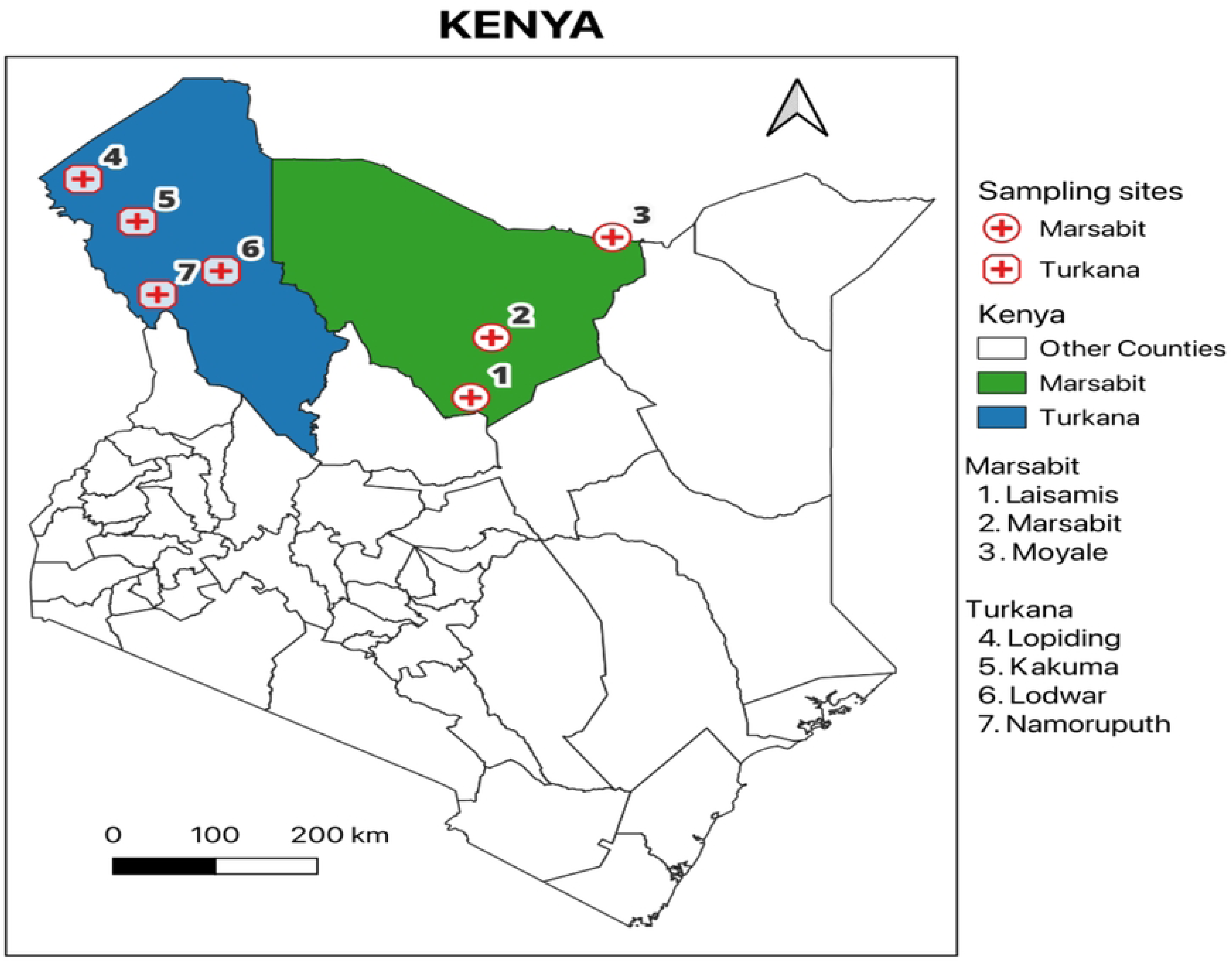
Map of Kenya developed using QGIS LTR (Version 3.40.14), showing the location of health facilities where the study was conducted.

### Study Procedures

Demographic and clinical data were collected and recorded from all patients who visited the treatment facilities and consented to participate in the study. The participants were tested using rK39 IT-Leish, rK28, and rK39 Kala-azar Detect, according to the manufacturers’ instructions. In line with national guidelines, rK39 IT-Leish was performed first, followed by rK28 and Kala-azar Detect. All tests were performed and interpreted by trained laboratory personnel using standardised operating procedures. Clinical management followed the national diagnostic guidelines (IT-Leish rK39 followed by DAT for case confirmation), ensuring that the results of study RDTs did not influence patient care (**Fig 2**).

**Fig 2:**
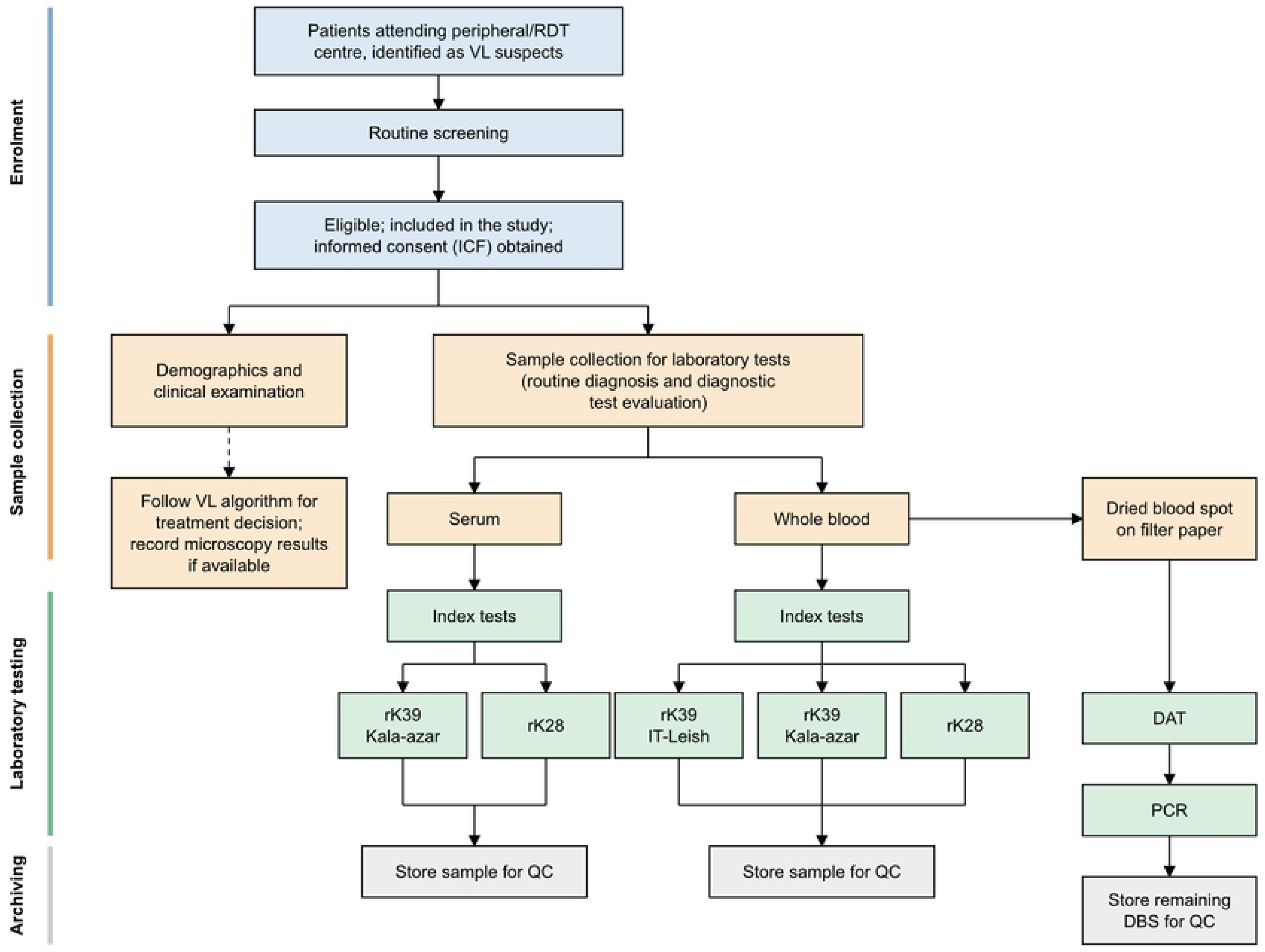
Recruitment of participants, sample collection and analysis plan.

**Figure 3:**
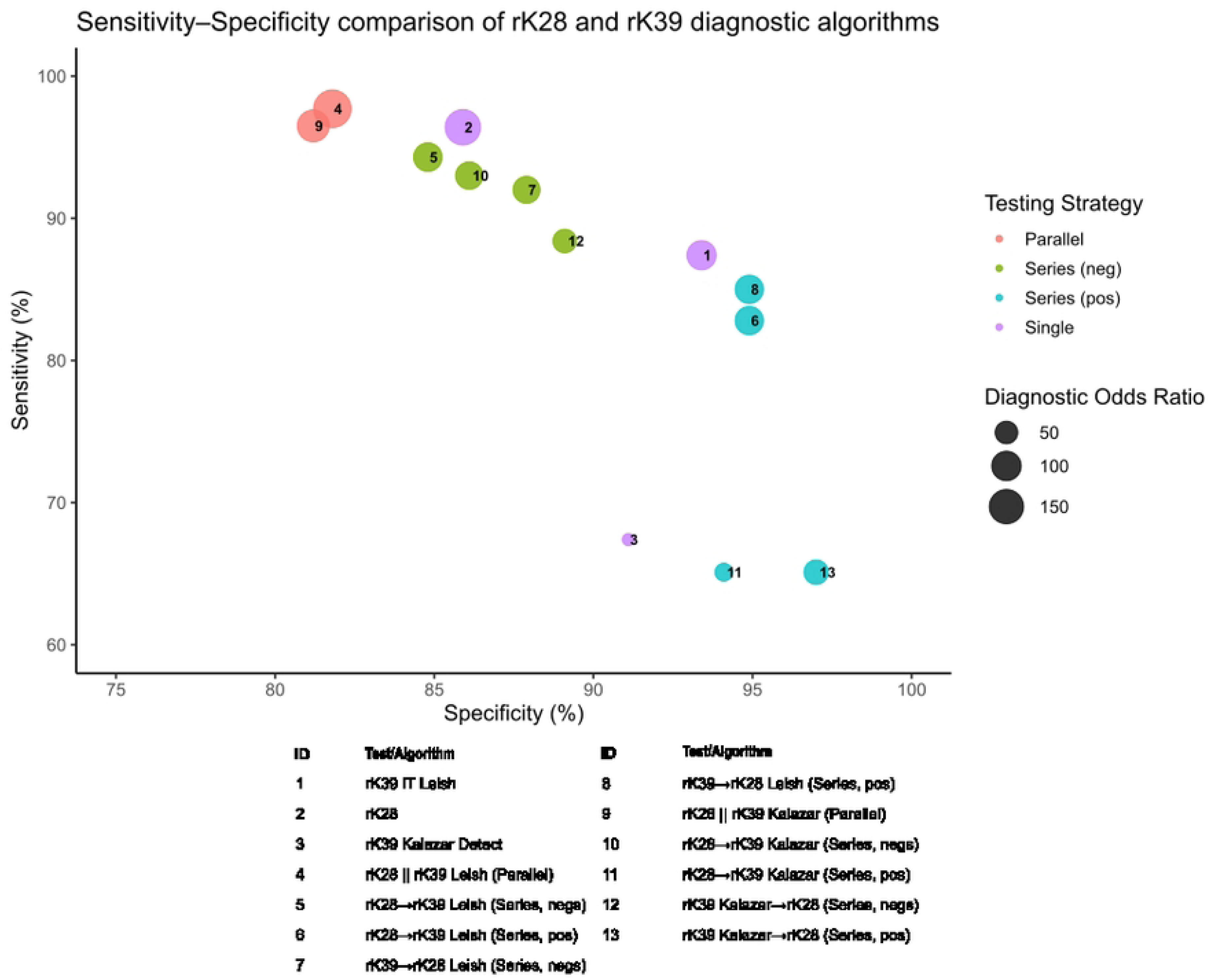
Comparative analysis of the Sensitivities, Specificities, and Diagnostic Odds Ratio of the different tests/algorithms.

For this study, DAT was used as the primary reference standard for diagnostic accuracy analyses. None of the health facilities sampled conducted microscopy as it is not a routine procedure, and there was limited clinical competence to undertake splenic aspirates and post-aspiration care. PCR results were excluded from the final analysis due to low DNA yields affecting assay performance.

### Interim Analysis

This analysis is based on the first 398 enrolled participants from a planned interim analysis at midterm recruitment, meeting the pre-specified minimum number of confirmed cases for primary accuracy endpoints. Full recruitment was not achieved due to operational constraints, including insecurity at the sites that interrupted enrolment, and disruptions in the national supply and subsequent discontinuation of the rK39 IT-Leish test by the manufacturer.

### Data Collection and Blinding

Data were recorded on standardized case report forms (CRFs) linked to unique study identification numbers. A structured questionnaire was administered to study participants to collect information on their sociodemographic characteristics, treatment-seeking experiences, and cost-related information. Demographic information collected included sex, age, location of origin, and occupation. Participants were also asked whether there were any delays in seeking healthcare and the reasons for the delays. Information on transportation costs incurred while seeking treatment was also collected. Index test results did not influence clinical decision-making for participants. Two laboratory technicians were assigned, with one responsible for reference tests and the other for index tests. The outcomes of the index test results were not blinded to the laboratory technicians who performed these tests. The IT-Leish rK39 was performed first, and the outcome was used to initiate patient management according to the national guidelines. Interpretation bias was minimised through standardised procedures, independent recording, and pictures(24). Pictures of RDT results were taken to enable source document verification and validation of results. The health facility location information (GPS Coordinates) was also collected for mapping (**Table 1**).

**Table 1:** Name and coordinates of health facilities in Marsabit and Turkana Counties from which participants were enrolled.

| **Site** | **County** | **Latitude** | **Longitude** |
|---|---|---|---|
| Laisamis Sub County Referral Hospital | Marsabit | 1.6168 | 37.8075 |
| Marsabit County Referral Hospital | Marsabit | 2.3299 | 37.9922 |
| Moyale County referral | Marsabit | 3.5231 | 39.0574 |
| Lopiding Sub-County Hospital | Turkana | 4.2149 | 34.3831 |
| Kakuma Missionary Hospital | Turkana | 3.7119 | 34.8614 |
| Lodwar County Referral | Turkana | 3.1225 | 35.6012 |
| Namoruputh (PAG) Health Centre | Turkana | 2.8414 | 35.0402 |

### Statistical Analysis

Diagnostic accuracy metrics, including sensitivity, specificity, positive and negative predictive values, diagnostic odds ratios, and balanced accuracy, were calculated for each test and algorithm for the combined parallel and series testing algorithms using DAT as the reference standard. Confidence intervals for proportions were estimated using the Wilson score method, while log-transformed values were applied for diagnostic odds ratios. Analyses were conducted for predefined populations based on completeness of index and reference test data. Parallel and serial algorithmic combinations were evaluated. In the parallel testing, a participant was classified as positive if any test was positive, with serial testing requiring concordant positive results across the tests. Sequential strategies involving retesting initial negatives or positives were similarly assessed. Analysis was done and plots generated using R-Studio (Version 2025.09.2 Build 418).

### Ethical Approval

Ethical approval was obtained from the Scientific Ethics Review Unit (SERU) at Kenya Medical Research Institute (KEMRI), under protocol number SERU 4436. A research permit No. NACOSTI/P/22/18928 was obtained from the Kenya National Commission for Science, Technology, and Innovation (NACOSTI). Regulatory approval was granted by the Kenya Pharmacy and Poisons Board for the importation and use of the index tests.

Written informed consents and assents were obtained from study participants before enrolment and from parents or legal guardians for participants below 18 years of age. All data were recorded on standardised case report forms (CRFs) and linked to unique study identification numbers. No personal identifiers were collected, ensuring participant confidentiality. Participants were informed that specimens would be used for VL diagnostic test evaluation.

## Results

A total of 398 participants were enrolled across the seven sentinel sites between January 2023 and March 2024. Laisamis (n=123) and Marsabit (n=88) contributed more than half of the participants (53%), reflecting historically high VL burdens at these sites. The study population was predominantly male (53.5%), and most participants (67.8%) were older than 15 years of age. Clinically, splenomegaly (20.4%) and hepatomegaly (12.6%) were the most common symptoms in addition to fever. HIV co-infection was uncommon, observed in only one participant from Laisamis (**Table 2**).

**Table 2:** Demographic and clinical characteristics of study participants. The rK28 RDT demonstrated a sensitivity of 96.4% (95% CI: 89.9–99.3%) and 85.9% (77.8– 91.9%) specificity. The rK39 IT-Leish RDT exhibited sensitivity of 87.4% (78.0–93.8%) and specificity of 93.4% (86.9–97.3%). By contrast, the Kala-azar Detect rK39 test showed the lowest sensitivity at 67.4% (56.0–77.5%43), although specificity remained high at 91.1% (83.6–96.0%) (**Figure 3**).

| **Characteristic** | **Laisamis (n=123)** | **Marsabit (n=88)** | **Lopiding (n=61)** | **Kakuma (n=46)** | **Lodwar (n=36)** | **Moyale (n=25)** | **Namoruputh (n=19)** | **Total (n=398)** |
|---|---|---|---|---|---|---|---|---|
| **Age group** |  |  |  |  |  |  |  |  |
| **<5 years** | 17 (13.8%) | 8 (9.1%) | 7 (11.5%) | 5 (10.9%) | 3 (8.3%) | 4 (16.0%) | 2 (10.5%) | 46 (11.6%) |
| **5–15 years** | 32 (26.0%) | 14 (15.9%) | 12 (19.7%) | 9 (19.6%) | 6 (16.7%) | 5 (20.0%) | 4 (21.1%) | 82 (20.6%) |
| **>15 years** | 74 (60.2%) | 66 (75.0%) | 42 (68.9%) | 32 (69.6%) | 27 (75.0%) | 16 (64.0%) | 13 (68.4%) | 270 (67.8%) |
| **Gender** |  |  |  |  |  |  |  |  |
| **Male** | 65 (52.8%) | 49 (55.7%) | 33 (54.1%) | 24 (52.2%) | 19 (52.8%) | 13 (52.0%) | 10 (52.6%) | 213 (53.5%) |
| **Female** | 58 (47.2%) | 39 (44.3%) | 28 (45.9%) | 22 (47.8%) | 17 (47.2%) | 12 (48.0%) | 9 (47.4%) | 185 (46.5%) |
| **Clinical signs** |  |  |  |  |  |  |  |  |
| **Fever** | 41 (33.3%) | 19 (21.6%) | 18 (29.5%) | 15 (32.6%) | 9 (25.0%) | 4 (16.0%) | 5 (26.3%) | 111 (27.9%) |
| **Splenomegaly** | 28 (22.8%) | 15 (17.0%) | 15 (24.6%) | 10 (21.7%) | 7 (19.4%) | 3 (12.0%) | 3 (15.8%) | 81 (20.4%) |
| **Hepatomegaly** | 19 (15.4%) | 9 (10.2%) | 9 (14.8%) | 5 (10.9%) | 4 (11.1%) | 2 (8.0%) | 2 (10.5%) | 50 (12.6%) |
| **Co-infections** |  |  |  |  |  |  |  |  |
| **HIV+** | 1 (0.8%) | 0 (0.0%) | 0 (0.0%) | 0 (0.0%) | 0 (0.0%) | 0 (0.0%) | 0 (0.0%) | 1 (0.3%) |

The rK28 kit had a higher sensitivity as compared to rK39 IT Leish, while specificity was higher in the rK39 as compared to rK28. A combination of the three kits in various algorithms (parallel, positive sequential or negative sequential) was employed to determine their effect on sensitivity, specificity and overall balanced accuracy.

Parallel diagnostic algorithms combining rK28 and rK39 IT-Leish offered the highest sensitivity, reaching 97.7%, although specificity decreased due to increased false positives. Sequential or series-based algorithms improved specificity, particularly when rK28 was used as the initial test, followed by rK39 IT-Leish for confirmation. Combinations involving rK28 and Kala-azar Detect performed better than Kala-azar Detect alone but remained inferior to rK28 paired with rK39 IT-Leish. Overall, rK28 consistently outperformed the other two RDTs, both individually and within algorithmic combinations (**Fig 4**).

**Fig 4:**
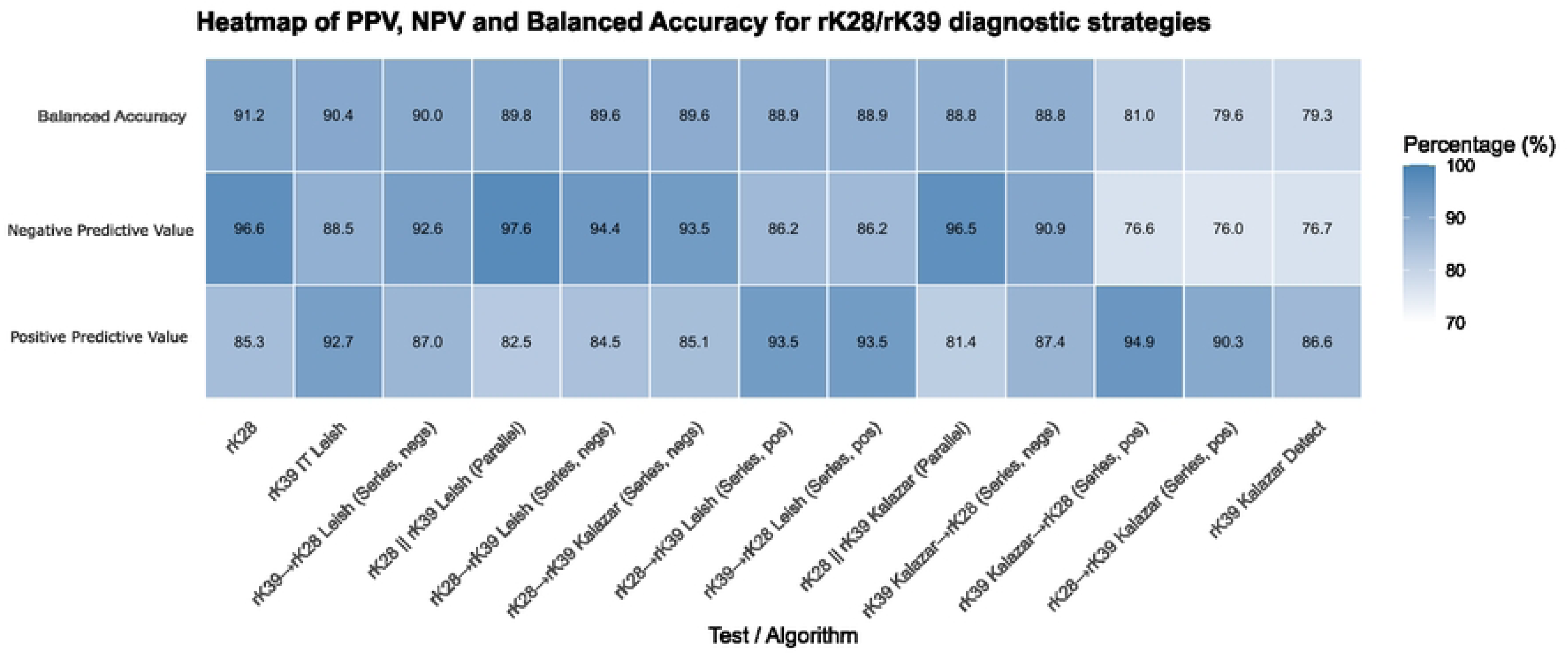
Comparative analysis of the Negative and Positive Predictive Values of the different tests and algorithms. The tests and algorithms are sorted by their balanced accuracy. The rK28 kit has the highest balanced accuracy but is more prone to false positives as compared to the rK39 IT Leish.

Socioeconomic factors related to diagnostic delay and incurred diagnostic costs were also analyzed. These factors were categorized into subgroups, namely, location, sex, age, HIV status, occupation and reason for delay in seeking healthcare.

The reported reasons for diagnostic delay were lack of financial resources (35.1%), lack of/inadequate transportation (22.9%), long distance to health facility (28%), unaware of where to access diagnosis (11.9%), Other (1.4%) and None (0.7%). Patients reporting no reason for their delay in seeking healthcare had the longest diagnostic delay, averaging 28 days. In comparison, those reporting reasons other than the four main ones had the shortest diagnostic delay, averaging 16 days **(Fig 5A).**

**Fig 5:**
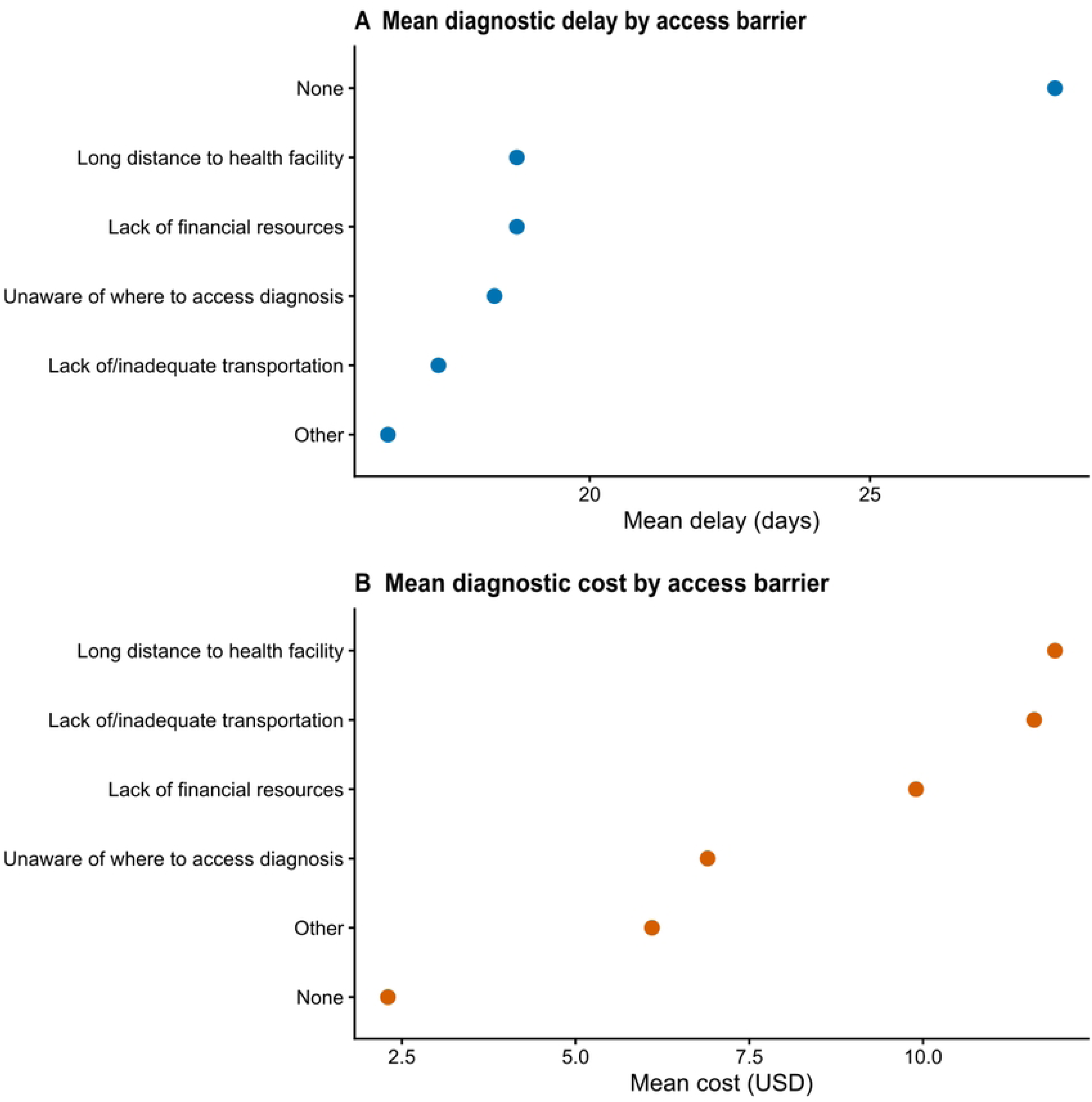
Comparison of the effects of access barriers to the mean diagnostic delay (A) and the mean cost of treatment (B). Distance to health facilities and transportation limitations had the biggest impacts on the cost of diagnostics. In contrast, those patients with no discernible barrier to seeking healthcare were the most likely to face a delay in getting a diagnosis.

In terms of impact on the incurred diagnostic costs, patients with reasons pertaining to distance and transportation to health facilities experienced the highest costs, averaging 11.38 USD and 11.40 USD, respectively. Patients reporting no reason to delay in seeking healthcare experienced the lowest cost, averaging 2.33 USD **(Fig 5B).**

Comparative analysis of mean patient delays and diagnostic costs across study locations revealed that patients from locations in Turkana County tended to exhibit higher diagnostic costs compared to those in Marsabit County. Namoruputh showed the highest average diagnostic cost at USD 21 with an average delay of 15.4 days, whereas Laisamis showed the least average diagnostic cost of USD 3.04 with an average delay of 19.5 days. In terms of patient delays, Lodwar had the longest average delay of 34.9 days, with an average diagnostic cost of USD 7.6 while Moyale and Lopiding reported less than 15 days of delay on average with the former also reporting less than USD 5 in diagnostic costs (**Fig 6**).

**Fig 6:**
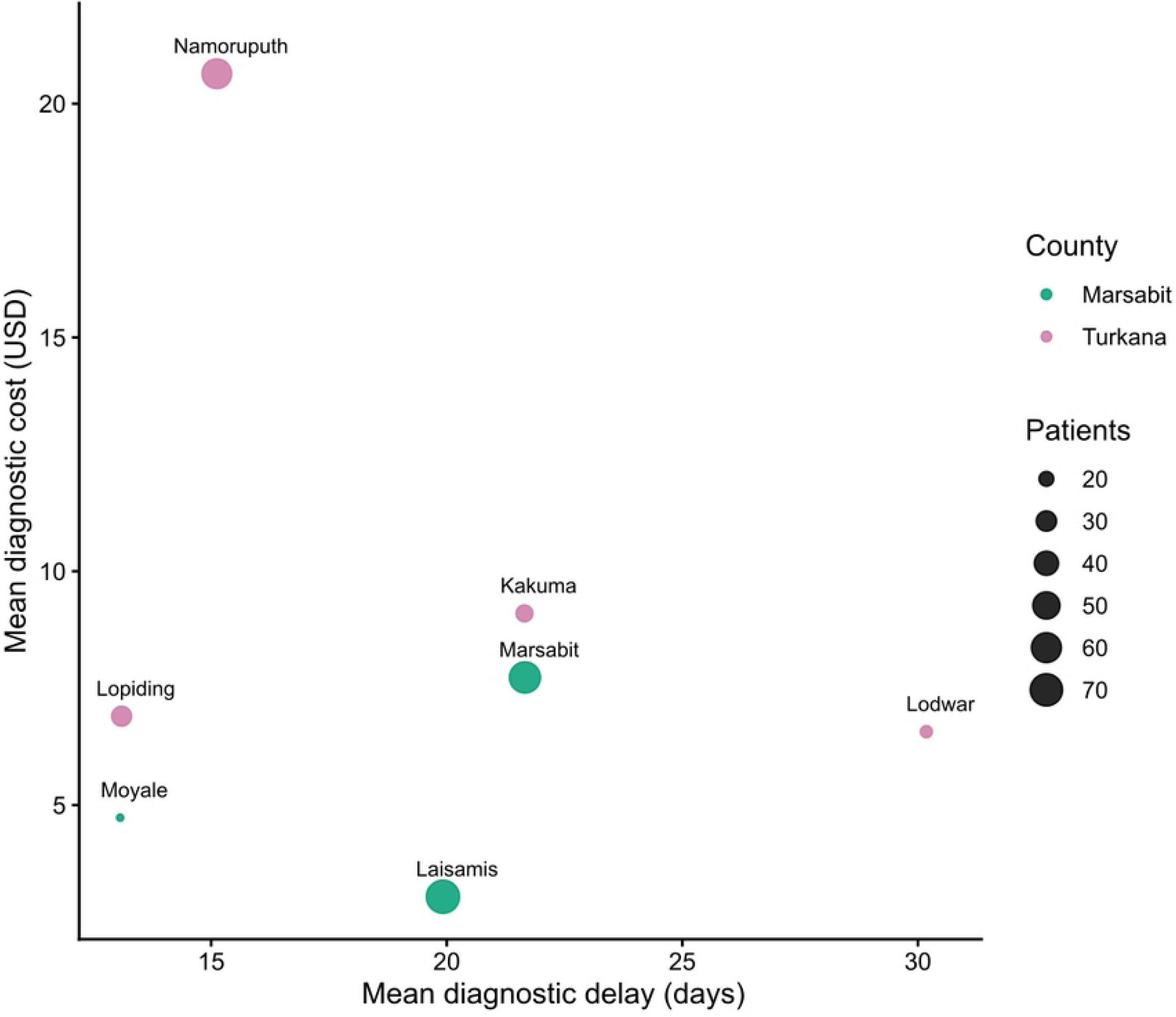
Comparison of the mean diagnostic delay and mean diagnostic cost as per county. Marsabit County had the highest number of patients, with most coming from Marsabit and Laisamis towns. Marsabit town had the highest diagnostic cost and patient delays as compared to Laisamis and Moyale. Namoruputh, in Turkana County, had the highest diagnostic cost incurred, exceeding all other towns in both counties by a significant margin. On the other hand, Lodwar had the longest diagnostic delays with an average delay of a month before patients sought out healthcare.

Comparison of delay and diagnostic costs across different patient groups showed that although male patients had longer delayed (with more variability) as compared to their female counterparts on average the treatment costs were nearly similar. When stratified by age, children under 5 years had a slightly lower delay but slightly higher diagnostic costs as compared to older patients. More variability in terms of distribution within a group was seen in diagnostic costs as compared to delay when occupation was considered. However, some occupations such as miners, farmers, refugees, and formal employees had little representation within the sampled population (**Fig 7**).

**Fig 7:**
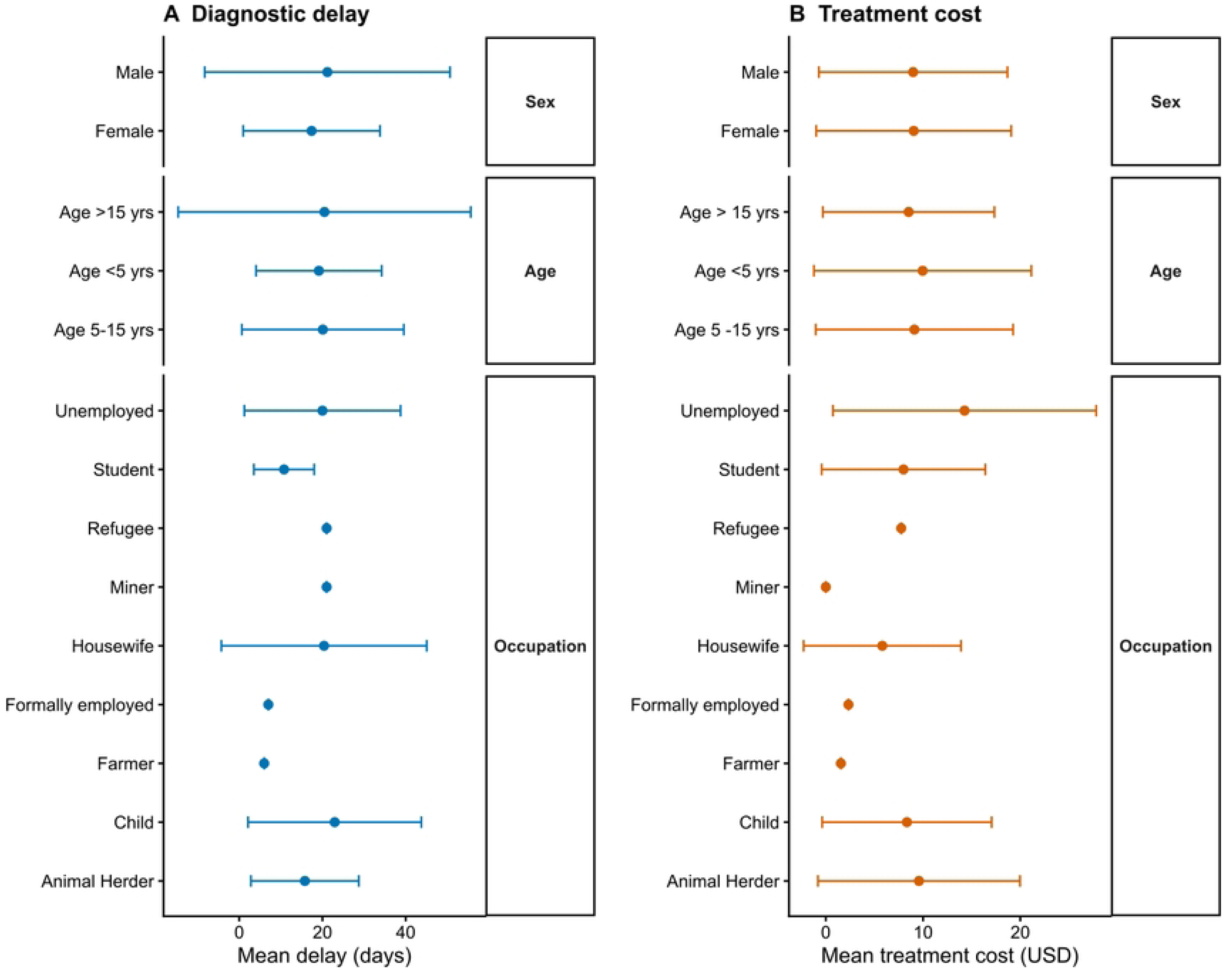
Comparison between delay in diagnosis (A) and cost of diagnosis (B) stratified by sex, age and occupation. The sex of the patient had little to no impact on the diagnostic cost as compared to diagnostic delay. Although male patients had a longer average delay, the delays also showed greater variability. Older age groups (5 years and above) were similar in diagnostic delays and diagnostic costs. Patients younger than 5 years experienced shorter diagnostic delays but slightly higher diagnostic costs. Occupation had the highest variability in terms of delay and treatment costs, although some occupational groups including refugees, miners, farmers and formally employed patients were underrepresented.

## Discussion

This study evaluated the diagnostic accuracy and operational characteristics of rK28 and rK39-based RDTs, as well as multiple algorithmic diagnostic approaches, for VL diagnosis in East Africa. Additionally, the study also investigated sociodemographic factors that may influence health-seeking behaviors, specifically looking at delays in going to the health facility and incurred diagnostic costs. The findings provide critical context to regional diagnostic needs amidst ongoing changes in test availability and programmatic priorities.

The rK28 RDT demonstrated a sensitivity of 96.4% in whole blood, consistent with Mukhtar et al. (2015)(36), who reported 92.5% sensitivity and 100% specificity in Sudan. Another study done by Mukhtar et al in 2018 in Sudan(37) reported a sensitivity of 96.7% and a specificity of 98.7% reaffirming the performance of the kit (37). These concordant results highlight rK28’s strong diagnostic potential across diverse East African settings. However, the specificity observed in this study was (85.9% [77.8–91.9]). Similarly, Cloots et al. (2020) reported a pooled specificity of 74.7% and sensitivity of 97.2% in Ethiopia and Sudan(38), illustrating the sensitivity–specificity trade-off observed with rK28-based RDTs. While ideal for initial screening, these tests require confirmatory diagnosis or clinical verification to reduce overtreatment risks. Operational advantages of rK28 include stability at high temperatures and a 24-month shelf life, addressing common challenges in storage and distribution. The frequent IT-Leish rK39 stockouts observed in this study further underscore the value of alternative platforms like rK28. However, its clinical adoption remains limited due to its research-use-only status, pending additional evidence, regulatory approval, and inclusion in national diagnostic guidelines.

The IT-Leish rK39 test, recommended by the WHO for VL diagnosis and incorporated into Kenya’s national diagnostic algorithm, demonstrated high specificity of 93.4% but comparatively lower sensitivity 87.4%. These findings align with earlier studies done by Mbui et al that reported similar low sensitivity in Kenya 89.3%(39) and regional meta-analyses by Cunningham et al, which reported a pooled sensitivity of 85.1% and specificity of 96.0%(40). Kala-azar Detect rK39 provides a high specificity of 91.1% and moderate positive predictive value of 86.6%, making it a reliable tool for ruling in VL, although a low sensitivity of 67.4% was observed in this study. This low sensitivity is consistent with findings from Ethiopia and Sudan(38,41), and its moderate diagnostic performance (balanced accuracy 79.3%), limit its suitability for elimination-focused programs. While the Kala-azar Detect rK39 test may serve as a pragmatic interim solution in low-volume or resource-constrained clinics, reliance on this single-test strategy risks missed cases and may undermine efforts toward interruption of VL transmission (42).

In this evaluation, both parallel and sequential (negatives retested) combinations of rK28 and rK39 Kala-azar Detect maintain high sensitivity (≥93.0%) and balanced accuracy (approximately 89%), comparable to those algorithms previously utilizing IT-Leish. The greatest sensitivity was observed with the parallel rK28‖rK39 Leish strategy at 97.7%, closely followed by parallel rK28‖rK39 Kala-azar Detect at 96.5%. Both approaches demonstrated high negative predictive values (NPV ≥96.5%), an essential attribute for screening initiatives in elimination programs, where the detection of all true cases is critical(33,43). Such findings are consistent with multicenter and modelling studies that show parallel algorithms significantly increased case detection and are advantageous in elimination programs where the risk of missing infected individuals must be minimized(38,41,44).

In VL elimination settings, prioritizing sensitivity and NPV is critical to avoid missed cases, even if specificity is moderate (81–86%)(45,46). Where drug toxicity remains a concern, positives-retested algorithms (e.g., rK28→rK39 Kala-azar Detect) may be used for confirmatory testing, offering higher specificity (94.1%) but lower sensitivity (65.1%). However, the improved safety profile of newer VL treatments like miltefosine(47), reduces the risks of overtreatment, supporting the use of high-sensitivity diagnostic algorithms to minimize missed cases and advance elimination efforts.

Double-test algorithms can enhance diagnostic accuracy for VL and support elimination goals in East Africa but involve higher costs, logistical complexity, and greater demands on health systems. Their success depends on formal inclusion in national guidelines, flexible policies, and strong support systems, including training, supply chains, and health information infrastructure. However, with coordinated planning, these strategies can turn improved accuracy into real progress toward VL elimination.

The findings of this study show that rK28 has a higher sensitivity and balanced accuracy compared to the rK39-based RDTs evaluated. These results suggest rK28 is particularly suitable in elimination settings for use in peripheral health facilities, where missing true VL cases have important clinical and epidemiological implications. The observed performance differences between rK28 and the Kala-azar Detect rK39 test are consistent with previous reports highlighting variability in the performance of rK39-based assays in East African settings (48).

The results align with previous evaluations conducted in Ethiopia, Sudan, and South Sudan, which have consistently demonstrated improved diagnostic performance of rK28 compared with rK39-based tests. The consistently high sensitivity of rK28 across regions suggests that it may be less affected by antigenic variability among circulating *L. donovani* strains. In contrast, the variable performance of rK39-based tests in East Africa may reflect both parasite diversity and host immunological factors.

Parallel testing algorithms further improved sensitivity, approaching near-complete case detection. This is particularly relevant for VL elimination efforts, where undiagnosed cases contribute to ongoing transmission. However, increased sensitivity comes with reduced specificity, which could lead to overtreatment in some contexts. Series testing approaches increased specificity, making them more suitable for confirmatory purposes or settings where overtreatment poses significant clinical or programmatic risks.

Collectively, the findings highlight the need for diagnostic strategies that are both high-performing and contextually feasible, particularly for underserved or high-risk populations. The results inform recommendations for health system strengthening and underscore the value of adapting interventions to local realities.

While the study achieved valuable insights into VL diagnostics, enrolment did not reach the initially planned sample size, which resulted in insufficient statistical power to perform secondary analyses such as cost-effectiveness assessment and evaluation of optimal test placement. Nonetheless, the data collected provided a robust basis for evaluating the primary diagnostic performance objectives. Second, according to the protocol, the reference standard comprised DAT, PCR on DBS specimens, or microscopy where available; however, no microscopy was performed due to facility constraints, and all PCR assays on DBS were negative, due to low DNA yields from the DBS. Consequently, analyses relied primarily on DAT as the reference, which may introduce misclassification bias owing to its imperfect sensitivity and specificity compared to parasitological or molecular confirmation.

## Conclusion

Algorithmic testing strategies and platforms such as rK28 present substantial opportunities to strengthen VL diagnostics and support elimination objectives in East Africa. However, the durable impact will depend on their adoption as clinical-use tests (rK28), incorporation into policy and national guidelines, and the underpinning of resilient supply chains and health systems. Optimisation of diagnostic strategies tailored to local capacity, epidemiology, and programmatic aims will be essential for maximizing accuracy, limiting overtreatment, and advancing regional VL elimination objectives. Continued operational evaluation remains critical to fully realise the potential of available RDTs in the evolving landscape. Notably, building on the diagnostic accuracy data presented, further research on the cost-effectiveness analyses is recommended to guide practical implementation and ensure efficient, sustainable progress toward VL elimination targets.

Given the discontinuation of IT-Leish and the lower performance of Kala-azar Detect rK39, Kenya’s national diagnostic algorithm should be revised to integrate rK28 as the primary diagnostic tool. Strengthening supply chains, ensuring wide availability of high-performing RDTs, and building capacity for their effective use will be essential for achieving national and regional VL elimination targets.

## Data Availability

All relevant data supporting the findings and conclusions of this paper are provided in the article. The raw data sets are publicly available and can be access here https://doi.org/10.6084/m9.figshare.34808034

